# Detection of human IgG antibodies against *Mycoplasma genitalium* using a recombinant MG075 antigen

**DOI:** 10.1101/2024.11.19.24317541

**Authors:** Anna Overgaard Kildemoes, Olivia Sureya Seierø Rai, Elisabeth Probst Lyng Westermann, Henrik Frederik Bekkevold Johansen, Eva Bjørnelius, Carin Anagrius, Liang Ma, Ida Rosenkrands, Jørgen Skov Jensen

**Affiliations:** Reproductive Microbiology, Department of Bacteria, Parasites & Fungi, Statens Serum Institut, Copenhagen, Denmark; Department of Dermatovenereology, Karolinska University Hospital Huddinge, Stockholm, Sweden; Department of Venereology, Falu lasarett, Falun, Sweden; Critical Care Medicine Department, NIH Clinical Center, National Institutes of Health, Bethesda, Maryland, United States of America; Infectious Disease Immunology, Center for vaccine research, Statens Serum Institut, Copenhagen, Denmark

## Abstract

*Mycoplasma genitalium* is a sexually transmitted pathogen, which can cause a range of reproductive tract diseases in both men and women. To disentangle the relationship between *M. genitalium* infection(s) and subsequent reproductive health complications at the population level, accurate serological tools are needed. The major challenge in developing specific *M. genitalium* serological tests is the extensive cross-reactivity with the closely related ubiquitous respiratory tract pathogen, *M. pneumoniae*. In this report, we describe the development of an immunoblot assay based on a recombinant fragment of the *M. genitalium* MG075 protein present in lipid-associated membrane extracts. A sensitivity of 87.1% was achieved based on testing antibody responses in sera from 101 adults with PCR-confirmed *M. genitalium* infection. A specificity of 95.2% was obtained through evaluation of sera from 166 children under 15 years of age with and without *M. pneumoniae* infection, who were unlikely to have been exposed to sexually transmitted *M. genitalium*. The development of a serological assay capable of accurately distinguishing *M. genitalium* and *M. pneumoniae*, will enable a better understanding of associations between *M. genitalium* and adverse reproductive sequelae.

## Introduction

*Mycoplasma (M.) genitalium* is a sexually transmitted bacterial pathogen, which can cause a wide range of mild to severe reproductive tract diseases in both men and women (1). Infections with *M. genitalium* have been linked to conditions such as urethritis, cervicitis, adverse pregnancy outcomes (including preterm birth and spontaneous abortion), and pelvic inflammatory disease (2).

Despite being first described in 1981, significant gaps in understanding the global epidemiology of *M. genitalium* persist (3). Molecular methods for the detection of current infections, evaluation of treatment efficacy and surveillance of antimicrobial resistance exist (4). However, these tools are not yet implemented as standard practice globally. The vast majority of population-level studies have been performed in high-income countries, reporting a one to two percent prevalence of *M. genitalium* infection in both women and men (5), while a meta-analyses including a limited number of studies from highly diverse low- and middle-income countries estimated a prevalence of 3.9% (6). To understand the risks and consequences of previous exposure(s) to *M. genitalium*, population-level studies based on highly specific antibody detection are needed. Previous studies focusing on the detection of *M. genitalium-*specific antibodies have relied primarily on whole cell or lipid-associated membrane protein (LAMP) antigen preparations from the G37 type-strain (7-10). Additionally, a few studies have used a mixture of LAMP antigens from two different strains to cover potential strain diversity (11, 12). Both whole-cell and LAMP antigen-based approaches mainly rely on detecting antibodies that recognize crude full-length antigen targets including the MG191 and MG192 proteins, which are part of the complex terminal structure involved in *M. genitalium* adhesion (1, 13). Two studies using crude G37 antigens demonstrated an association between the presence of anti-MgPa antibodies and tubal factor infertility (14, 15). The results from non-standardized assays across different studies pose significant challenges for comparison and interpretation in epidemiological research. Furthermore, all current serological tests for *M. genitalium* suffer from considerable cross-reactivity with the common respiratory pathogen *M. pneumoniae* (16, 17). The cross-reactivity is attributed to the presence of orthologs for all proteins encoded in the highly compacted genome of *M. genitalium* (580kb) within the slightly larger genome of *M. pneumoniae* (816kb) (18). Therefore, it is desirable to develop an assay that targets defined epitopes unique to *M. genitalium* with no or minimal cross-reactivity with *M. pneumoniae* to achieve high specificity. A highly specific serological test would be crucial for disentangling the associations between prior exposure to *M. genitalium* and subsequent reproductive health complications at the population level. Here we present the performance of a recombinant protein fragment (MG075F1) of the *M. genitalium* MG075 gene in an immunoblot assay. The MG075 protein was chosen because our preliminary investigation of immunoreactive targets in LAMP preparations gave a positive signal in serum samples from *M. genitalium* infected individuals on immunoblots (Suppl. Figure S1). Mass spectrometry confirmed that this reactivity was caused by the sparsely expressed MG075 protein (Suppl. Figure S2B). Apart from proteins highly expressed from the MgPa operon, this is one of the few *M. genitalium* antigens recognised by IgG antibodies in *M. genitalium-*positive people. We took the advantage of access to sera from adults with PCR-confirmed *M. genitalium* infection to determine the sensitivity of the MG075F1 antigen for detecting IgG antibodies. Specificity was determined using samples from children under 15 years of age, both with and without serologically confirmed *M. pneumoniae* infection.

## Methods

### *M. genitalium* MG075F1 cloning, expression, and purification

Details about cloning, production and purification of the recombinant *M. genitalium* MG075F1 protein fragment in *Escherichia coli* are provided in Supplementary Materials. Briefly, the coding region for the N-terminal 798 amino acids of MG075 (MG075F1) was amplified using primers designed based on the *M. genitalium* G37 strain sequence (GenBank accession no. L43967.2) and used to produce recombinant protein with the pET102 TOPO® vector and expression system (Invitrogen K102-01). The N-terminal 798 amino acids were selected because the immediate next codon, TGA, codes for tryptophan in mycoplasmas species, while functioning as a stop codon in *E. coli* used to express the recombinant protein. The *MG075F1* plasmid was transformed into BL21-(DE3) *E. coli* by electroporation, and expression of the recombinant protein fragment was confirmed by Western blotting using crude induced cell lysate and anti-His-tag antibody. Since, the recombinant MG075F1 protein formed inclusion bodies, purification was done under denaturing conditions. Fractions with a high content of MG075F1 (∼107kDa including thioredoxin and tags, see Suppl.) and a low content of non-target protein were chosen (Suppl. Figure S3). The optimal amount of antigen for line-Western blotting was determined through antigen titration using known positive and negative sera as well as anti-His tag antibody.

### Amino-acid sequence BLAST and alignments

A standard protein blastp search on GenBank was done using the protein sequence including the first 798 amino acids of MG075 predicted from the genome of the *M. genitalium* G37 type strain (GenBank accession no. L43967.2). Additional MG075 protein sequences were retrieved from full genome assemblies of other *M. genitalium* strains (19) and compared using the CLC Genomics Workbench version 20.0.

### SDS-PAGE and immuno-blotting

In the optimized assay, 1 µg MG075F1 antigen in 1x Laemmli (Biorad #1610737), 50mM DTT, 8M urea was separated on 4-20% SDS-PAGE gels (precast mini-protean TGX, 7cm single well, Biorad #4561091) at 100V constant voltage for 50 min in standard 25 mM Tris, 192 mM glycine, 0.1 % SDS running buffer. Antigen was transferred using trans-blot turbo mini 0.2 µM nitrocellulose transfer packs (Biorad #1704158) at 25V for 3 min/gel in a Trans-Blot Turbo Transfer System (Biorad #1704150). Membranes were blocked for 30-60 minutes at room temperature and then over-night at 4°C in Tris-buffered saline with Tween (TBST, Medicago #097510) with 5% skimmed milk powder (blocking/sample buffer). Sera were diluted 1:200 in sample buffer and 600µl loaded onto blots using a Mini-Protean II line-blotter with a capacity for 20 samples per blot (Biorad #170-4017) and incubated on a shaker for one hour. A standardised aliquot of a pool of known positive sera was used as positive control on all blots. Blots were washed thrice in in TBST for five minutes on a shaker after removal from the line-blotter. Incubation with alkaline phosphatase (AP) conjugated y-chain specific goat anti-human IgG (Sigma-Aldrich #A3150 1:4000) secondary antibody was done in sample buffer on a shaker for 1 hour. Blots were washed three times for five minutes on a shaker in TBST before 10ml 1-step BCIP/NBT substrate (ThermoScientific #34042) was added. Development was stopped with Milli-Q washes and blots dried before imaging and analyses. All incubations except O/N blocking were done at room temperature.

### Protein band image analysis

White light images of dried blots were obtained using the GBOX Chemi-XRQ system and analysed using GeneTools (version 4.3.17.0) software. The manual band quantification setting with normalization to an internal known positive control on the same blot was done for each sample line. Local background was subtracted for each individual sample line and all blots were manually inspected for artefacts. An image analysis cut-off on relative signal normalized to the internal standardized positive control (≥25) combined with mean pixel unit (≥700) was used as criteria for positive signal. This cut-off was determined based on samples with and without PCR confirmed *M. genitalium* infection as well as samples from children unlikely to ever have had an *M. genitalium* infection. Fischer’s exact test with a significance level of 0.05 was used to compare nominal variables. Data visualization was made with GraphPad Prism (10.2.2).

### Serum sample sets

For evaluation of the specificity of MG075F1, sera from Danish children (1-15 years of age) with a history of respiratory symptoms with and without complement fixation test antibodies against *M. pneumoniae* (MPT) (20) were selected as they were unlikely to have been exposed to sexually transmitted *M. genitalium*. The serum samples comprised 62 MPT positive and 104 MPT negative. Titers ≥64 were considered positive. The use of these anonymized sera for diagnostic method developments was not subject to ethical committee approval due to the anonymized nature of the dataset. Sensitivity was determined using sera from Swedish adults (n=101) with *M. genitalium* PCR-positive urogenital samples. One cohort sample set (Sample set 1: n=76 individuals with follow-up available for n=30) was collected in 2002-2004 with ethical approval from the Uppsala Research Ethical Committee (journal no. 2010/429). A second sample set (Sample set 2: n=25 individuals with follow-up samples available for n=19) was collected in 2003-2004 with ethical approval for the use of these sera for *M. genitalium* serology granted by the ethics committee in Stockholm, Sweden, D.nr: 392/01 (21). No data about the sex of the patients was available for sample set 2. For both sample sets 1 and 2 the first serum sample available was obtained one to four weeks after the positive *M. genitalium* PCR (the date on the first available sample is set to day 1 in this study).

## Results

### Strain variation of MG075F1

The sequence of the MG075F1 protein fragment from *M. genitalium*, which consists of the N-terminal 798 amino acids from MG075 was highly conserved as demonstrated by the alignment of 18 unique MG075 sequences from 29 clinical isolates collected between 1980 and 2010 from Australia, Japan, and five different European countries (19). For the MG075F1 region, these 29 isolates had 99.4-100% amino acid identity whereas the pairwise similarity with *M. pneumoniae* ranged from 52.1-52.5% (Figure 1 and suppl. Figure S4). A blastp (protein-protein BLAST®) search revealed that 13 *M. pneumoniae* sequences were only 51.6-52.1% identical (96-100% query coverage). These *M. pneumoniae* sequences were all from the P116 (MPN213) lipid acquisition surface protein. Other identified P116 family protein sequences with low degree of similarity belonged to known human mollicutes as *M. pirum* (23.2%; 74.3% coverage) and *M. amphoriforme* A39 (25.7%; 69% coverage) as well as a penguin and a tortoise associated species; *M. tullyi* (23.6%; 71% coverage) and *M. testudinus* (25.5%, 80% coverage). No putative MG075F1 homologs were found in the blast search in other mollicutes like *Ureaplasma* sp., *Acholeplasma* sp., and other *Mycoplasma* species such as *M. hominis*. The high conservation of the MG075F1 sequence across *M. genitalium* isolates, combined with its significant divergence (∼50%) from the *M. pneumoniae* P116 sequences (Figure 1), underlines the potential of this protein fragment as a valuable target for differential serology.

**Figure 1:**
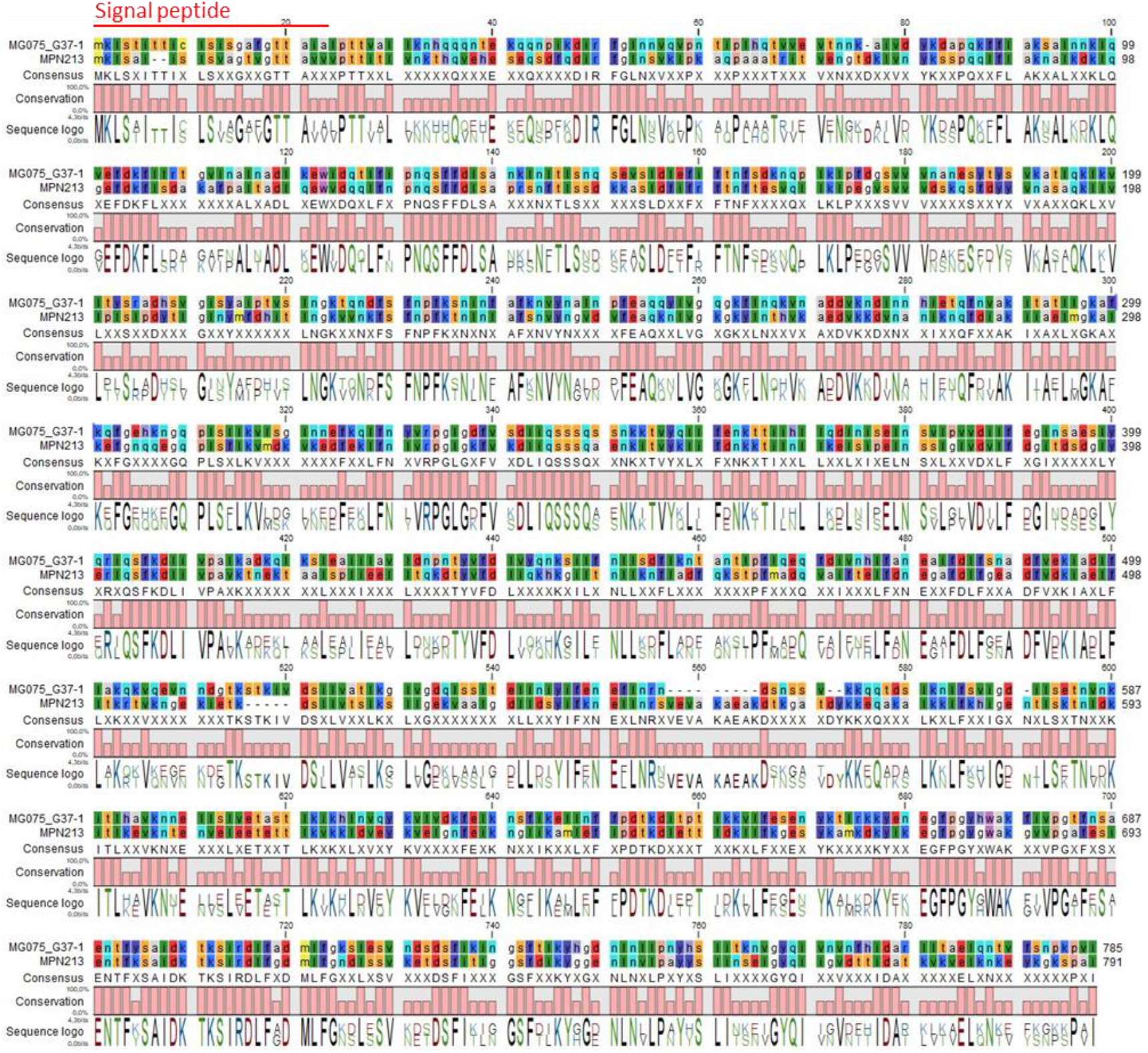
Sequence alignment of MG075F1 from *M. genitalium* and the corresponding P116 (MPN213) fragment from *M. pneumoniae*. MG075: G37 GenBank full genome accession no. L43967.2 and predicted protein accession no. AAC71293.1. *M. pneumoniae* MPN213: NCBI accession no. WP_010874570. The predicted signal peptide is indicated with a red horizontal line.

### Sensitivity and specificity of MG075F1 in the IgG line-blot assay

IgG binding to recombinant MG075F1 was assessed on Western blots in a line-blot format, which allowed assessment of 17 samples and three controls per blot (Figure 2). Through image analyses with normalization to a standardized positive control, specificity and sensitivity was determined. The overall specificity using samples from children (n = 166) was 95.2% (95% CI: 91-98%) with an image analysis cut-off <25 of relative signal normalized to the positive control combined with mean pixel unit <700 (Table 1). There was no difference in the proportion of samples containing cross-reactive IgG MG075F1 antibodies between the 104 MPT-negative children and the 62 MPT-positive children (Fischer’s exact test = 1). Furthermore, no association between MPT titer and the presence of IgG against MG075F1 was seen for the MPT-positive samples.

**Table 1:** Specificity of MG075F1 based on *M. pneumoniae* complement fixation test positive and negative samples from children (≤15 years old) stratified by age groups and MPT titer.

| SPECIFICITY | MPT dilution factor | Age in years (range: 1-15 - mean: 9.4 - median: 10) |  |  |  |  |
| --- | --- | --- | --- | --- | --- | --- |
|  |  | 1 - 15 | 1 - 4 | 5 - 8 | 9 - 12 | 13 - 15 |
| MPT negatives | $\leq 16$ | 94.2% (98/104) | 100% (25/25) | 93.3% (14/15) | 90.0% (27/30) | 94.1% (32/34) |
| MPT positives | $\geq 64$ | 95.2% (59/62) | 100% (4/4) | 93.3% (14/15) | 96.2% (25/26) | 94.1% (16/17) |
|  | 64 | 3 neg |  | 1 neg | 1 neg | 1 neg |
|  | 128 | 9 neg |  | 1 neg | 3 neg | 5 neg |
|  | 256 | 4 neg | 1 neg |  | 2 neg | 1 neg |
|  | 1024 | 3 pos, 30 neg | 3 neg | 1 pos, 8 neg | 1 pos, 13 neg | 1 pos, 6 neg |
|  | 2048 | 8 neg |  | 3 neg | 3 neg | 2 neg |
|  | 4096 | 5 neg |  | 1 neg | 3 neg | 1 neg |
| **TOTAL (%)** ($n_{\text{neg}}/n_{\text{total}}$) | | 95.2% (158/166) | 100% (29/29) | 93.3% (28/30) | 92.9% (52/56) | 94.1% (48/51) |

MPT = *Mycoplasma pneumoniae* test (IgG). MPT is considered positive $\geq 64$ . IgG anti-Mg075F1 pos=positive and neg=negative.

**Figure 2:**
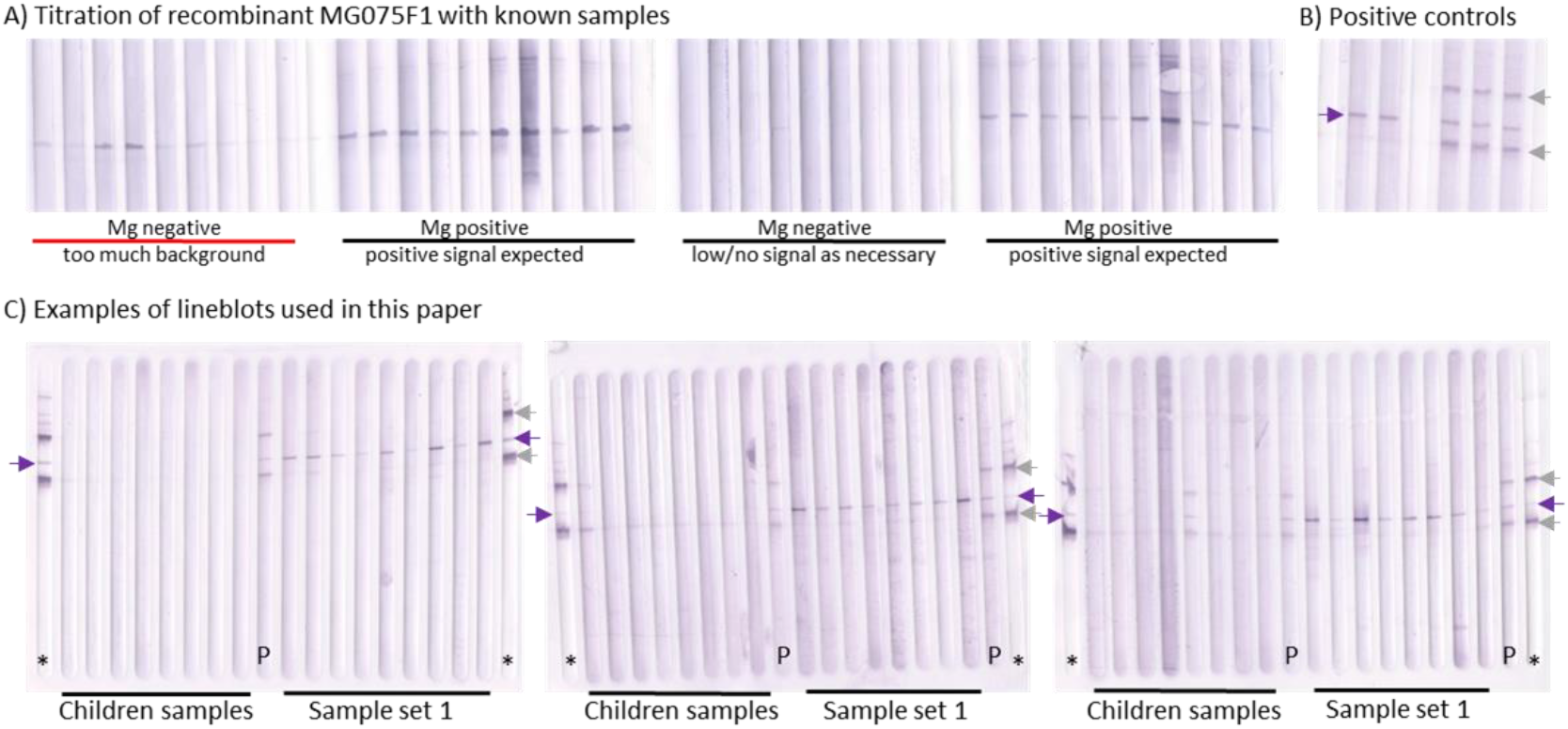
Optimisation of the MG075F1 IgG Western lineblot assay and examples sample incubations. A) Illustrates an example of titration of purified recombinant MG075F1 with known samples. The two lineblots were run with different amounts of antigen and incubated with identical sample material from known Mg negative and Mg positive individuals. The purple arrow indicates MG075F1 antigen. B) Shows an example of standardised positive controls. This blot was run with a combination of MG075F1 antigen (purple arrows) and antigen (grey arrows) not presented in this paper. This demonstrates the potential for multiplexing this lineblot assay. Left most is a duplicate of the positive control pool made from only known Mg positive sera and right most a triplicate of the same Mg positive control pool with an addition of known sera reactive with antigens not presented in this paper (grey arrows). C) Three examples of lineblots incubated with, respectively, samples from children (Mp±) and sample set 1. Purple arrows indicate MG075F1 antigen. The position of anti-His-tag controls (*) and positive pool sera controls (P) are indicated on each blot. Controls were loaded on each side of the blot and in the middle to ensure that alignment of the relevant antigen was always identifiable. Image analysis was done on the GBOX Chemi-XRQ system and GeneTools (version 4.3.17.0) software. A-C are blot scans made with a desktop image scanner.

Sensitivity was assessed based on two sample sets from 101 adults with PCR-confirmed *M. genitalium* infection. The overall sensitivity of MG075F1 was 87.1% (95% CI: 79-93%) based on the latest collected sample available for each individual (Table 2 and Figure 3C). Sensitivity was 86.8% (66/76) in sample set 1 and shown broken down stratified by sex and per time point in table 2. No data on sex was available for the second sample set from *M. genitalium* PCR positive adults (Figure 2B). The sensitivity of MG075F1 in the second sample set was 87.5% at day 1 (Time 1; 21/24) and 88.0% (22/25) when the last available sample was used (Time 2, 3, or 4). To get an indication of the longevity of IgG responses to MG075F1, the available subset of follow-up samples for both sample sets was measured (Figure 2 B, C). In general, once IgG specific for MG075F1 was detectable it was sustained within the time frames available. The longest sero-persistence demonstrated here is 533 days where the first visit sample was positive. One individual was IgG positive 616 days after the first sample was provided, however, this sample had an IgG level under the cut-off for the first visit, so it is not possible to determine the exact duration of IgG positivity.

**Table 2:** Sensitivity of MG075F1 based on *M. genitalium* PCR positive samples from adults stratified by sex and clinic visit (Sample set 1)

| SENSITIVITY | Total* | Time 1 | Time 2 | Time 1 only<sup>o</sup> | Time 1 paired<sup>+</sup> | Time 2 paired<sup>+</sup> |
| --- | --- | --- | --- | --- | --- | --- |
| **Sample set 1 (%)**<br>($n_{\text{pos}}/n_{\text{total}}$) | 86.8% (66/76) | 81.4% (57/70) | 88.9% (32/36) | 85.0% (34/40) | 76.7% (23/30) | 86.7% (26/30) |
| male | 85.4% (35/41) | 83.8% (31/37) | 85.7% (18/21) | 85.0% (17/20) | 82.4% (14/17) | 82.4% (14/17) |
| female | 88.6% (31/35) | 78.8% (26/33) | 93.3% (14/15) | 85.0% (17/20) | 69.2% (9/13) | 92.3% (12/13) |

\*Combination of Time 1 and Time 2 data to include all available individuals. If paired samples were available Time 2 was used (includes six Time 2 samples with no Time 1 sample available). <sup>o</sup>No Time 2 samples available. <sup>+</sup>Paired Time 1 and Time 2 samples as shown in Figure 2 for sample set 1.

**Figure 3:**
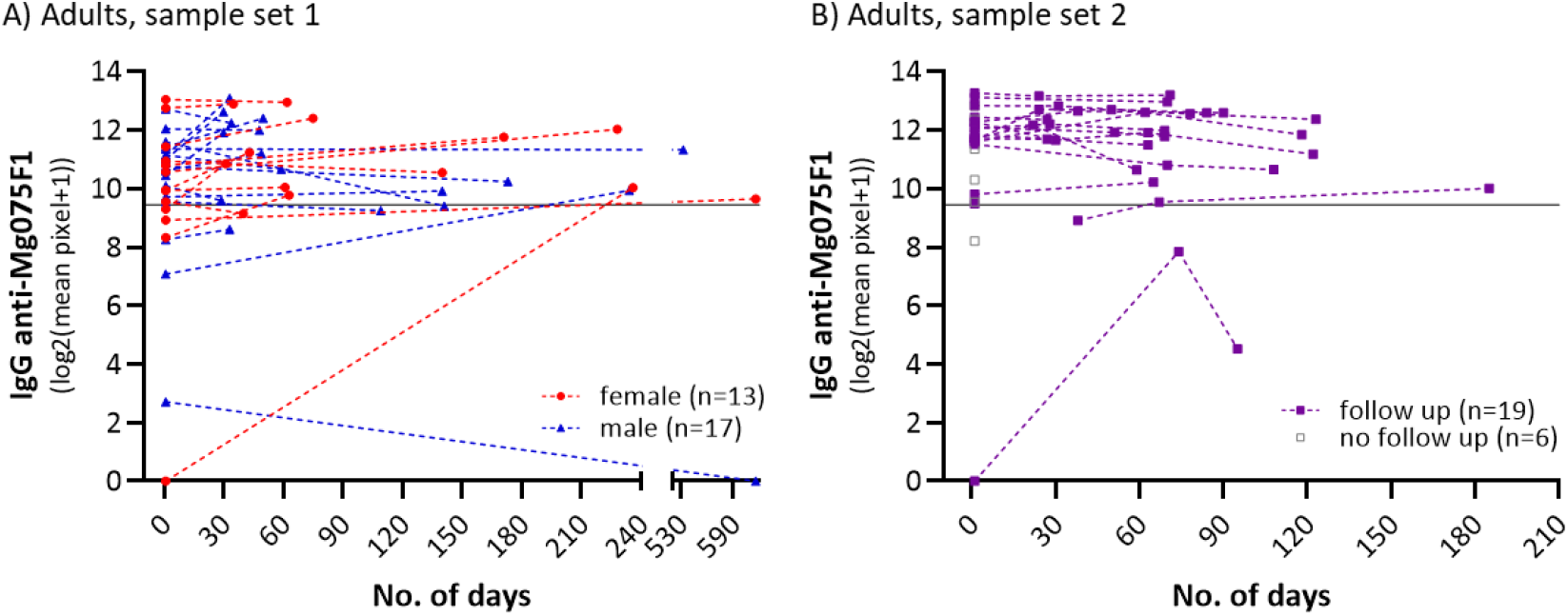
IgG anti-MG075F1 responses in samples from *M. genitalium* PCR positive adults. Longevity of IgG anti-MG075F1 responses shown in A) for sample set 1 consisting of individuals with paired samples available (n=30, table 2). Females are indicated with red circles and males in dark blue triangles. IgG anti-MG075F1 levels over time from sample set 2 are shown on B) in purple squares (n=19) and grey squares (n=6, no follow-up). Dotted lines are used to connect samples that are from the same individual over time (days from the first available sample) on A) and B) and the pixel signal cut-off value is indicated with a horizontal line.

## Discussion

The major challenge of accurately determining *M. genitalium* exposure (prior and/or current) based on serological assays is the high frequency of cross-reactive antibodies elicited through infection of the ubiquitous respiratory tract pathogen *M. pneumoniae* (16). Specificity is key to achieving a high predictive value of a positive test. However, for common infections caused by closely related pathogens with a high likelihood of shared epitopes, finding specific antigens is not trivial. During the last decade, sequencing of genomes from different *M. pneumoniae* and *M. genitalium* isolates has increased our knowledge about the conservation of putative antigens between these two species, highlighting the risk for cross-reactivity (19, 23). The antibody responses to *M. pneumoniae* and *M. genitalium* infections in humans are centered around a limited number of proteins with the surface-localized fraction receiving the most attention. For *M. genitalium*, adhesins expressed from the MgPa operon, including MG191 and MG192, have been identified as the main antibody targets in prior serological work (15, 24). These proteins have also been shown to generate specific antibody responses in non-human primate and rodent models (17, 25, 26). However, significant rapid antigenic variation driven by homologous recombination between genomic repeat regions in two of the genes contained in the MgPa operon (*mgpB* (MG191) and *mgpC* (MG192)) has been observed within isolates (27-29). This antigenic variation is thought to contribute to immune evasion and persistence of *M. genitalium* infection and thus, the specific antibody repertoire elicited in an individual may not be reactive against protein fragments from the strain selected for preparation of antigen. For a diagnostic assay such antigenic variation can compromise sensitivity and is expected to be an issue both for native and recombinant antigens.

In contrast to MG191 and MG192, MG075 is an attractive target, given its low strain-to-strain variation and presumed lack of recombination. Furthermore, the significant divergence from its counterpart in *M. pneumoniae* rendering the likelihood for cross-reactivity low. MG075 is most likely also surface-expressed, similar to its homolog in *M. pneumoniae*, P116 (MPN_213), which is known to be involved in essential lipid uptake (30). The interest in this antigen originated from the analysis of *M. genitalium* LAMP antigens, one of their components was found to be immuno-reactive with sera from *M. genitalium* infected patients and was subsequently identified as MG075 by mass spectrometry (Suppl. Figure S1 and S2). The present study confirmed, that anti-MG075F1 antibodies are useful biomarkers for discrimination between *M. genitalium* and *M. pneumoniae* infection based on IgG detection with a specificity of 95.2% and sensitivity of 87.1%. The observed specificity of 95.2% for IgG anti-MG075F1 was based on samples from Danish children 1-15 years of age, who were unlikely to have been exposed to or infected with sexually transmitted *M. genitalium*. It is not known, however, if children can become infected or colonized during birth which could explain the relatively high proportion of children with anti-MG075F1 reactivity. The likelihood of exposure to *M. pneumoniae* infection increases with age, thus, specificity might be expected to decrease in older children. However, we did not observe an increase in the frequency of false positives in the older groups of included children irrespective of their MPT status. This observation underlines the potential of MG075F1 as a discriminatory biomarker for serological differentiation of prior and/or current *M. genitalium* infection from *M. pneumoniae* exposures. The ubiquitous presence of IgG specific for multiple *M. pneumoniae* antigens has been shown in clinically healthy children from the age of seven years, and the frequency of IgG specific for more than one target increased with age (31). Several infections during childhood are likely required before robust anti-*M. pneumoniae* IgG responses are induced. It is undetermined whether a single *M. genitalium* infection is enough to generate IgG responses. However, due to the large repertoire of shared epitopes present in these respiratory and genital tract pathogens, prior *M. pneumoniae* infection(s) may enable rapid and robust initiation of later-life IgG response to *M. genitalium* infection. Our results demonstrate a persistent presence of IgG specific for MG075F1 up to at least 533 days after seroconversion. In general, once seroconversion has taken place, the observed IgG levels appeared stable in our sample set. Despite the small sample size, this observation is very encouraging in terms of using anti-MG075F1 in epidemiological studies to investigate the potential link between prior *M. genitalium* exposures with later-life reproductive health complications. Future studies should be designed with longer follow-up intervals and larger sample sizes to determine, how long the sensitivity of detection of IgG anti-MG075F1 is sustained. Interestingly, IgG specific for MG191 and MG192 in urethral swabs but not sera from men with non-gonococcal urethritis and PCR confirmed *M. genitalium* infection has been shown to generally decline (32). Similarly, IgG specific for the *M. genitalium* adhesins in cervical swabs has been shown to be transient, whereas the specific IgG levels in sera were sustained in a macaque infection model (26). It is unknown whether the local genital mucosal surface levels of IgG anti-MG075F1 also decline despite the presence of *M. genitalium* infection, however, it is likely that assays based on sera will perform better than those based on genital tract swabs.

This study was based on Western blots of recombinant MG075F1 antigen. This approach offers an advantage over whole-cell preparations, which suffer from batch-to-batch variation and have a higher cross-reactive potential due to the large number of epitopes, some of which might be shared with other pathogens or commensals. Furthermore, MG075F1 is a superior target compared to proteins expressed from the MgPa operon as much lower strain variability occurs at this site. One limitation of the recombinant MG075F1 is its highly hydrophobic nature, which leads to the formation of inclusion bodies. Consequently, the addition of 6-8 M urea to the loading buffer was necessary prior to gel electrophoresis if the protein had been stored at -20°C after purification. In general, the His-tag on the expressed construct was not readily available for detection with anti-his antibodies if the material was not kept under harsh denaturing conditions. Therefore, it is recommended to verify antigen presence with known positive controls in addition to traditional anti-his antibody detection. Future developments of diagnostic assays based on MG075F1 detection would benefit from protein engineering to increase the solubility of the protein. Epitope mapping could be used to identify essential and non-essential stretches of sequence within the MG075F1 protein, potentially allowing the use of this target under non-denaturing conditions and further improving assay specificity. Such protein engineering would pave the way for the use of other assay formats such as ELISA, bead-based assays and lateral flow formats. Incorporation of (part of) the highly specific MG075F1 into a bead-based platform is an attractive option as this supports multiplexing strategies for surveillance and epidemiological studies in combination with other relevant pathogens.

In conclusion, we demonstrate that MG075F1 is a highly specific antibody target for the differentiation of *M. genitalium* from *M. pneumoniae* exposure in an immunoblot assay format. This tool will be beneficial for elucidating the role of *M. genitalium* infection in reproductive health issues at the population level.

## Data Availability

All data produced in the present study are available upon reasonable request to the authors

## Acknowledgements

We would like to thank Vivi Andersen and Christina Nørgaard for excellent technical assistance with protein purification and cloning, respectively. We also thank Dr. David Martin and Dr. Miriam Mancuso (LSU Health Sciences Center, New Orleans, USA) for their assistance and support in initial identification of MG075. Parts of the present work were funded by an NIH grant to Brandie Taylor (1R01AI143653-01A1) with JSJ as sub-PI, an NIH/NIAID grant to Lisa Manhart (R01 AI61019) with JSJ as subcontractor. Furthermore, this work was supported in part by the grant: Gulf South Sexually Transmitted Infections/Topical Microbicide Cooperative Research Center (NIAID grant no. U19 AI061972) and the Intramural Research Program of the NIH Clinical Center, Bethesda, Maryland, USA.

