## Supplementary material for "Detection of human IgG antibodies against *Mycoplasma genitalium* using a recombinant MG075 antigen": Suppl fig S1-4

**Identification of MG075 in lipid-associated membrane protein (LAMP) preparations**

Lipid-associated membrane proteins (LAMPs) were prepared from the *M. genitalium* G37 type strain as previously described (1, 2) and separated on precast 4–12% Tris-glycine gradient gels (1.0 mm thick, Invitrogen) under reducing conditions. The proteins were transferred to Invitrolon PVDF (Invitrogen) at 20V for 2 hr, followed by incubation in Western Blocker solution (Sigma) at room temperature for at least 30 min. Immunoblotting was carried out using human sera diluted 1:200. Horseradish peroxidase (HRP)–labeled polyclonal antibodies against human IgG diluted 1:2,000 were used. Sera from *M. genitalium* NAAT positive patients reacted strongly with an approximately 110 kD antigen whereas sera from *M. pneumoniae* complement fixation test (MPT) positive patients were negative as were most sera from MPT negative children (Figure S1). In order to identify the 110 kD antigen, four LAMP preparations present in the hydrophobic phase made under the same conditions and the total cell lysate of the G37 type strain were resolved on an 8% Tris-glycine SDS-PAGE gel (1.5 mm thick, Invitrogen) under reducing conditions. The gel was stained with SYPRO Ruby protein stain (Molecular Probes, Invitrogen) and the bands of approximately 110 kDa from the four LAMP preparations were excised and pooled. Following trypsin digestion, the peptides were subjected to mass spectrometry (MS). MS analysis showed that the ~110 kDa band contained protein sequence matching MG075 (score 232) as identified by peptide query on the Mascot server (Matrix Science Ltd) (Figure S2B).

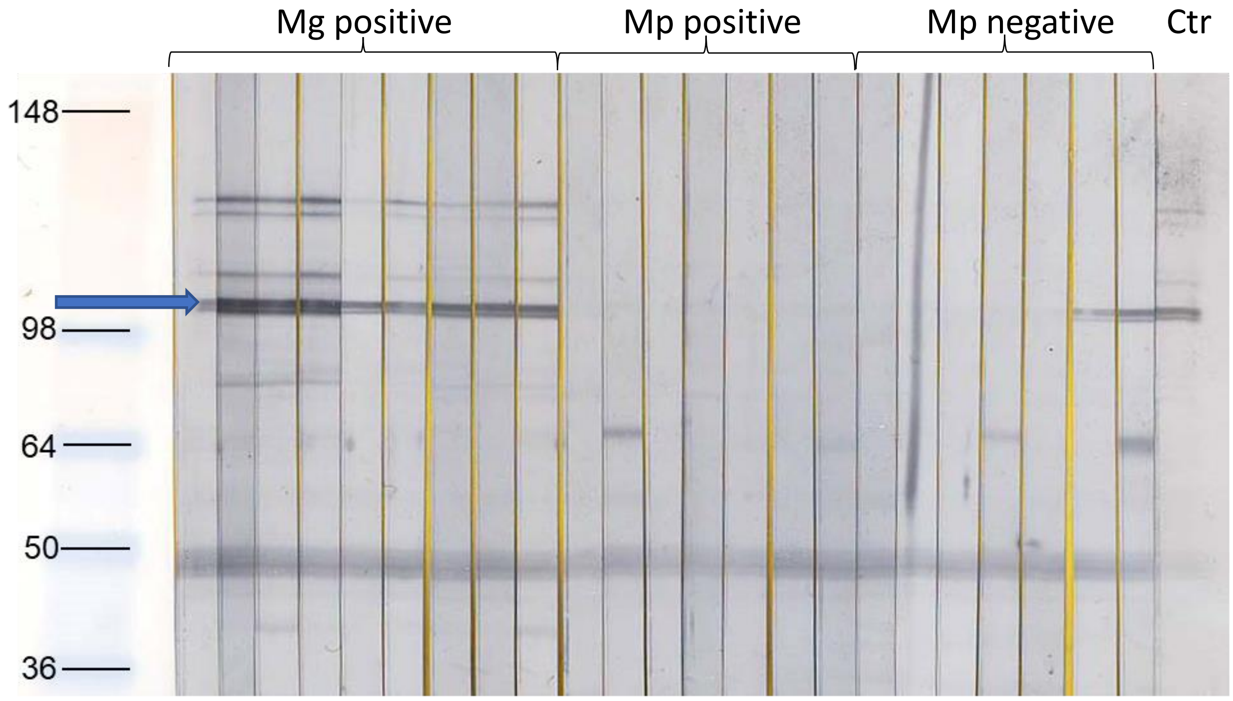


**Suppl. Figure S1**: Western blot of sera from *M. genitalium* NAAT positive patients (Mg positive) reacting with an approximately 110 kD antigen (blue arrow) whereas sera from *M. pneumoniae* complement fixation test (MPT) positive patients (Mp positive) were negative as were most sera from MPT negative children (Mp negative). On the right most side a pool of known Mg positive sera was used as positive control. The Mp positive and negative samples are from Danish children as described in the main manuscript. These Mg positive samples were from Louisiana State University, United States of America, where written informed consent was obtained from participants and the study protocol was approved by the Institutional Review Board of the Louisiana State University Health Sciences Center.

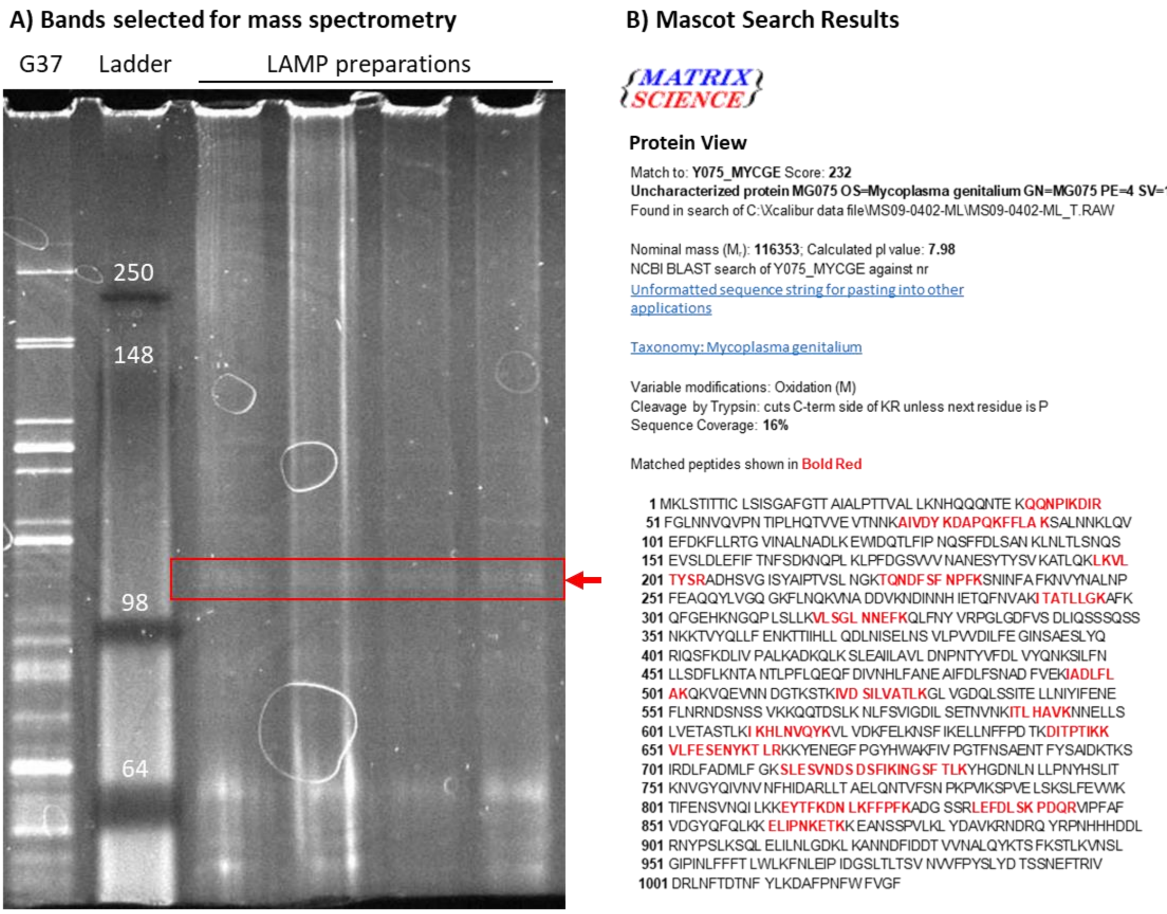


**Suppl. Figure S2: Confirmation of presence of MG075 in LAMP preparations by gel electrophoresis and mass spectrometry**

A) SYPRO Ruby protein stained SDS-PAGE gel with the following loaded (left to right): Total strain G37 protein preparation, reference protein ladder (band size in kDa given in white just above bands), four LAMP preparations. Box and arrow indicate the bands cut out and analysed by mass spectrometry. B) Mascot server search output confirming presence of MG075 in the bands cut from the gel. Note that A) is best visualized if opened on devices with (O)LED or other high contrast displays.

### *M. genitalium* MG075F1 cloning, production and purification

The N-terminal MG075F1 fragment was amplified using primers designed based on the *M. genitalium* G37 strain (accession no. L43967.2) for use in conjunction with the pET 102 TOPO® vector and expression system (Thermo Fisher Scientific, Invitrogen K102-01). Forward primer sequence was 5'-CACCATGAAATTAAGTACAATAACTACTATC-3' (cloning tag underlined) and reverse primer 5'-AACTTCAAATAAGGATTTGGAGAGTTCAAC-3'. As the usual stop codon TGA codes for tryptophan in mycoplasma species, the fragment length of 798 amino acids was determined by the first TGA occurrence in the reference sequence. DNA extracted from the Danish version of the *M. genitalium* G37 strain (3) was used as template for standard PCR amplification using the Platinum high fidelity *Taq* polymerase (Invitrogen). PCR products were purified (Qiagen QIAquick®, Qiagen, Hilden, Germany) and cloned into the plasmid vector with C-terminal 6x His-tag and carrying ampicillin as selection marker according to manufacturer's protocol (Invitrogen K102-01). After plasmid amplification in competent Top 10 *E. coli* and purification, the correct vector construct was confirmed by Sanger sequencing. The full amino acid sequence of the MG075F1 construct (~107kDa, isoelectric point ~6) including expressed vector components is as follows:

MGSDKIIHLTDDSFDTDVLKADGAILVDFWAHWCGPCKMIAPILDEIADEYQGKLTVAKLNIDHNPGTAPKYGIRGIPTLL
LFKNGEVAATKVGALSKGQLKEFLDANLAGSGSGDDDDKLGIDPFTMKLSTITTICLSISGAFGTTAIALPTTVALLKNHQ
QQNTEKQQNPIKDIRFGLNNVQVPNTIPLHQTVVEVTNNKAIVDYKDAPQKFFLAKSALNNKLQVEFDKFLLRTGVINALN
ADLKEWIDQTLFIPNQSFFDLSANKLNLTLSNQSEVSLDLEFIFTNFSDKNQPLKLPFDGSVVVNANESYTYSVKATLQKL
KVLTYSRADHSVGISYAIPTVSLNGKTQNDFSFNPFKSNINFAFKNVYNALNPFEAQQYLVGQGKFLNQKVNADDVKNDIN
NHIETQFNVAKITATLLGKAFKQFGEHKNGQPLSLLKVLSGLNNEFKQLFNYVRPGLGDFVSDLIQSSSQSSNKKTVYQLL
FENKTTIIHLLQDLNISELNSVLPVVDILFEGINSAESLYQRIQSFKDLIVPALKADKQLKSLEAIILAVLDNPNTYVFDL
VYQNKSILFNLLSDFLKNTANTLPFLQEQFDIVNHLFANEAIFDLFSNADFVEKIADLFLAKQKVQEVNNDGTKSTKIVDS
ILVATLKGLVGDQLSSITELLNIYIFENEFLNRNDSNSSVKKQQTDSLKNLFSVIGDILSETNVNKITLHAVKNNELLSLV
ETASTLKIKHLNVQYKVLVDKFELKNSFIKELLNFFPDTKDITPTIKKVLFESENYKTLRKKYENEGFPGYHWAKFIVPGT
FNSAENTFYSAIDKTKSIRDLFADMLFGKSLESVNDSDSFIKINGSFTLKYHGDNLNLLPNYHSLITKNVGYQIVNVNFHI
DARLLTAELQNTVFSNPKPVIKSPVELSKSLFEVKGELKLEGKPIPNPLLGLDSTRTGHHHHHH

**Colour code: Thioredoxin, enterokinase site, MG075F1, V5 epitope, His tag, Spacer**

The MG075F1 plasmid was transformed into BL21-(DE3) *E. coli* by electroporation and cultured under 0,1mg/ml ampicillin pressure in LB medium (Sigma-Aldrich L3397). Protein fragments were produced by induction of the BL21-(DE3) MG075F1 strain with 0.5 mM IPTG (OmniPur, Sigma-Aldrich 5810, Merck, Søborg, Denmark) at OD ~0,5 culture inoculation. After ~4 hours of growth, cells were harvested by centrifugation at 2500 *x g* 4$^{o}$C for 10 min and stored at -20°C temporarily. Successful induction of MG075F1 was confirmed by western blotting of crude boiling lysate material as described under SDS-PAGE and immunoblotting using 4-20% precast TGX-gels (BioRad, Copenhagen, Denmark; #4561091) with anti-his-AP (1:2500, Thermo Fisher Scientific, Novex R932-25) as detection antibody. Pellets were resuspended in ice-cold lysis-buffer of 400 mM NaCl, 50 mM Tris-HCL, 1% Triton X-100 (Merck, 108603) with EDTA-free protease inhibitors (Roche, Vallensbæk, Denmark, 4693159001) and transferred to tubes with 0,1-0,15mm zirconia-silicate beads (Biospec Products, Inc. Bartlesville, OK, USA; #110709101z). Mechanical lysis was done on a Tissuelyser II (Qiagen) for 3 min at 30 Hz twice and kept on ice for five minutes in between runs. Subsequently, material was pooled again and sonicated for 3 min with 4 sec on and 3 sec off at 57% output on a Sonoplus probe sonicator (Bandelin, Berlin, Germany). After a brief sedimentation of the zirconium beads, the supernatant was then centrifuged in new tubes at 22000 x *g* at 4$^{o}$C for 30 min. The expressed MG075F1 forms inclusion bodies, so the pellets were washed twice in the lysis buffer as above and once without Triton X-100 and then resuspended in 6 M GuHCl, 50 mM $Na_2HPO_4$, 400 mM NaCl (pH 8.0). The MG075F1 in buffer was sterile filtered before it was loaded onto a HisTrap™HP nickel column (1 ml Cytiva 17524701, Merck) on an NGC™ medium pressure liquid chromatography system (BioRad, with ChromLab version 6.0.0.35) with a fraction collector. The column was then washed in 8 M urea (Merck 108487), 100 mM $Na_2HPO_4$, 10 mM Tris-HCl (pH 8.0) and elution performed on a gradient by introduction of 8 M urea, 50 mM $Na_2HPO_4$, 500 mM imidazole (pH 8.0) and 1 ml fractions collected. Fractions with MG075F1 were identified by western blotting using 15-well precast gels (BioRad 4568096) with anti-his-AP as detection antibody. Fractions with positive anti-his signal were pooled and dialysed against 6 M urea, 20 mM Tris-HCl (pH 8.0) over a 10 kD MWCO snakeskin membrane (ThermoScientific 88245). Anion exchange was subsequently performed on a HiTrap™Q HP column (1 ml, Cytiva 17115301) with gradient elution by introduction of 6 M urea, 20 mM Tris-HCl, 1 M NaCl (pH 8.0) and collection of fractions. Location of MG075F1 in fractions was confirmed by anti-his-AP western blotting. Presence of non-target protein was determined by western blotting (polyclonal rabbit-anti-*E. coli* 1:1000 BioRad 4329-4906; goat-anti-rabbit-IgG-AP 1:3000 BioRad 170-6518) and standard silver-staining of gels. Fractions with high content of MG075F1 (107 kDa) and low content of non-target protein were chosen (see suppl. figure 1). Total protein concentrations in chosen fractions were estimated using a QUBiT protein assay (Invitrogen Q33211) and a QUBIT fluorometer (Invitrogen) according to manufacturer's instructions. A pool of the chosen fractions had a final estimated concentration of 0.092 µg/µl protein and contained predominantly MG075F1. A total of 1 µg protein was determined as suitable for line-blotting through antigen titration in combination with incubation of known positive and negative sera and anti-his stain controls.

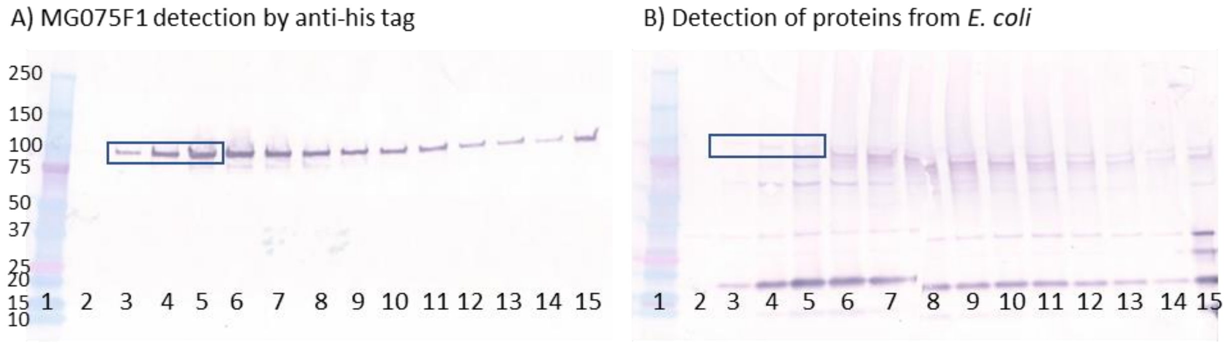


**Suppl. Figure S3: Representative Western blots of protein fractions collected after anion exchange**
Fractions loaded into lane 3, 4, and 5 (boxed) as shown on western blots for A) anti-his tag (MG075F1 107 kDa) and B) crude anti-*E. coli* proteins were chosen. These fractions had suitably low content of non-target protein with molecular weight just under the required 107kDa present in fractions from lane six and higher on B). Lane 1 contains a 10-250 kDa ladder (BioRad 161-0374) as specified on A). Lane 15 contains a pre-dialysis and anion exchange control.

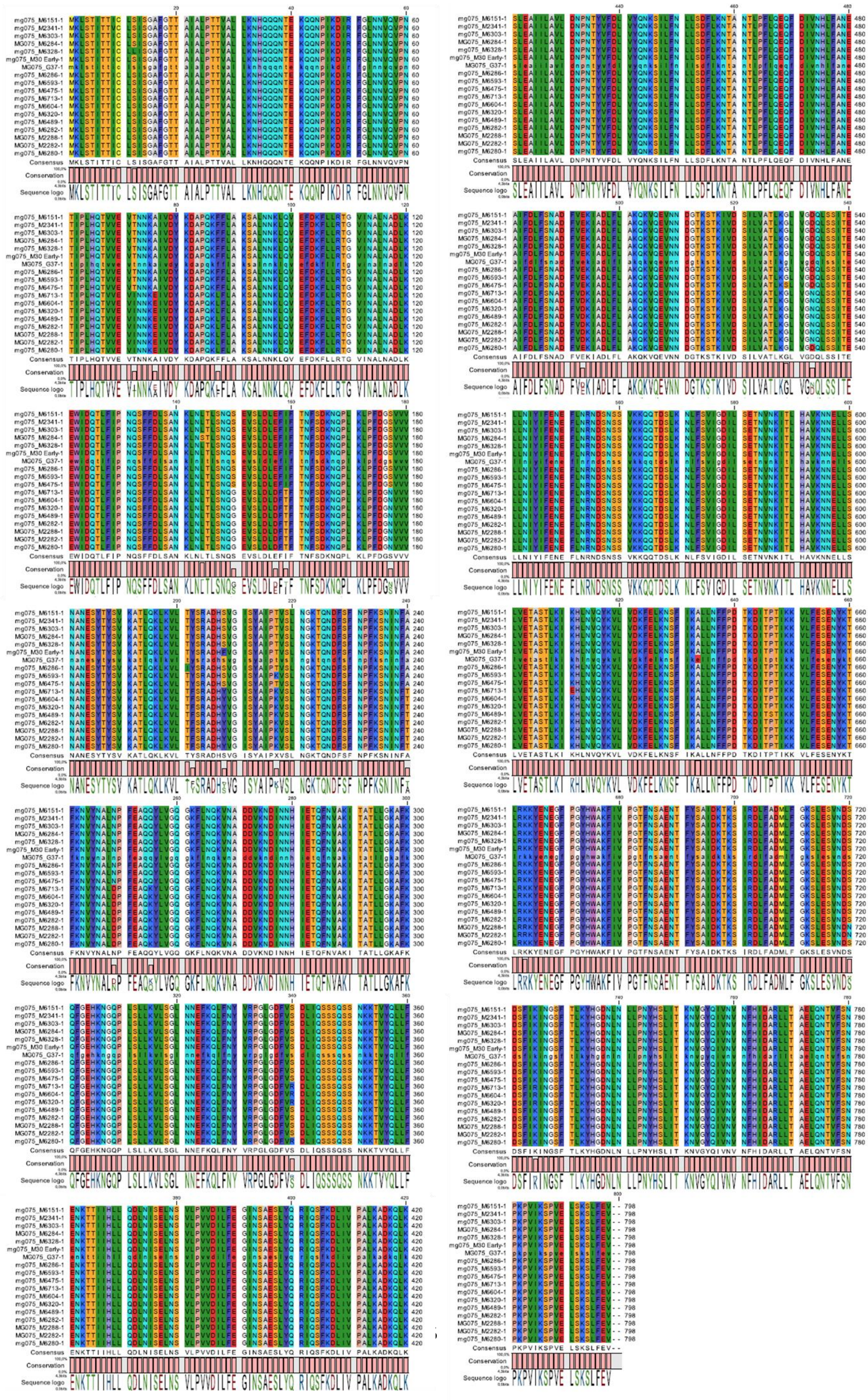

**Suppl. Figure S4: MG075F1 is highly conserved between *M. genitalium* strains**

The vector containing MG075F1 constructed in this paper is based on the G37 Danish isolate. Here alignment to 17 other unique *M. genitalium* MG075F1 sequences from isolates collected between 1980 and 2010 and representing five European countries, Japan and Australia are shown. These sequences as well as full genome sequence from ten other isolates with non-unique MG075F1 sequences can be extracted from the additional material included with Fookes *et al.* 2017 (3). Alignment was done using CLC Genomics Workbench ver. 20.0.
